# Absolute and relative excess mortality across demographic and clinical subgroups during the COVID-19 pandemic: an individual-level cohort study from a nationwide healthcare system of US Veterans

**DOI:** 10.1101/2023.05.12.23289900

**Authors:** Daniel M. Weinberger, Krishnan Bhaskaran, Caroline Korves, Brian P. Lucas, Jesse A. Columbo, Anita Vashi, Louise Davies, Amy C. Justice, Christopher T. Rentsch

## Abstract

**Background:** Most analyses of excess mortality during the COVID-19 pandemic have employed aggregate data. Individual-level data from the largest integrated healthcare system in the US may enhance understanding of excess mortality.

**Methods:** We performed an observational cohort study following patients receiving care from the Department of Veterans Affairs (VA) between 1 March 2018 and 28 February 2022. We estimated excess mortality on an absolute scale (i.e., excess mortality rates, number of excess deaths), and a relative scale by measuring the hazard ratio (HR) for mortality comparing pandemic and pre-pandemic periods, overall, and within demographic and clinical subgroups. Comorbidity burden and frailty were measured using the Charlson Comorbidity Index and Veterans Aging Cohort Study Index, respectively.

**Results:** Of 5,905,747 patients, median age was 65.8 years and 91% were men. Overall, the excess mortality rate was 10.0 deaths/1000 person-years (PY), with a total of 103,164 excess deaths and pandemic HR of 1.25 (95% CI 1.25-1.26). Excess mortality rates were highest among the most frail patients (52.0/1000 PY) and those with the highest comorbidity burden (16.3/1000 PY). However, the largest relative mortality increases were observed among the least frail (HR 1.31, 95% CI 1.30-1.32) and those with the lowest comorbidity burden (HR 1.44, 95% CI 1.43-1.46).

**Conclusions:** Individual-level data offered crucial clinical and operational insights into US excess mortality patterns during the COVID-19 pandemic. Notable differences emerged among clinical risk groups, emphasising the need for reporting excess mortality in both absolute and relative terms to inform resource allocation in future outbreaks.

**KEY MESSAGES:**

1. Most analyses of excess mortality during the COVID-19 pandemic have focused on evaluations of aggregate data, which may miss important individual-level drivers of excess mortality that may serve as future targets for improvement initiatives.
2. Using individual-level data from a national integrated healthcare system, we estimated absolute and relative excess mortality and number of excess deaths overall and within demographic and clinical subgroups.
3. Absolute rates of excess mortality were typically highest in groups where the baseline rate of mortality was higher; namely in older age groups and among those with more comorbidities and higher levels of physiologic frailty.
4. Relative measures of excess mortality were typically greatest among younger age groups and among those with lower physiologic frailty and fewer comorbidities.
5. Relative measures of excess mortality attenuated but remained elevated after censoring follow-up at first documented SARS-CoV-2 infection or COVID-19, suggesting that factors beyond SARS-CoV-2 infection contributed to the observed excess mortality during the pandemic.

## INTRODUCTION

During the COVID-19 pandemic, there was a substantial increase in rates of death due to any cause.^1–3^ Rates of deaths that exceed expected levels are referred to as excess deaths, which were observed globally.^4–6^ Some geographic regions, risk groups, and age groups experienced larger excesses,^4,6^ much of which was directly attributed to the virus, particularly in older adults.^7^ Other evidence points to healthcare system-level factors, such as disruptions to healthcare system function, personal health management, and healthcare utilisation.^8,9^ However, the pandemic also caused major disruptions in society, possibly contributing to overdoses,^10^ suicides,^8^ or violent crime.^11^ The risk of death due to COVID-19 as well as susceptibility to these secondary effects of the pandemic depends on a complex set of factors including the underlying health status of an individual.

To fully understand the drivers of excess deaths during the COVID-19 pandemic, including those caused directly by the virus and those indirectly caused by pandemic disruptions, it is necessary to consider detailed individual-level characteristics. Most analyses of excess deaths during the COVID-19 pandemic focused on evaluations of aggregate data, looking at changes in numbers of deaths compared to a pre-pandemic baseline. Linking these time series with data on other characteristics and risk factors can provide a broader understanding of the drivers of excess deaths.^5,7^ However, even this strategy may miss important individual-level drivers of excess mortality that may serve as future targets for improvement initiatives. With individual-level data from an integrated care system, it is possible to address this gap in knowledge and obtain a better understanding of the individual demographic and clinical factors that influence excess mortality and to identify the patient subgroups that experienced the greatest burden of excess deaths. Using individual-level data from the largest integrated healthcare system in the US, we estimated excess mortality rates and number of deaths overall and within demographic, comorbidity, and physiologic frailty subgroups. These analyses provide a more comprehensive picture of excess mortality than can be obtained from aggregate data alone.

## METHODS

### Data source

The US Department of Veterans Affairs (VA) serves 9 million Veterans annually at 171 medical centres and 1,112 outpatient sites nationwide.^12^ All care is recorded in an electronic health record with daily uploads into the VA Corporate Data Warehouse. Available data include demographics, outpatient and inpatient encounters, diagnoses, laboratory measures, and death records.

This study was approved by the institutional review boards of VA Connecticut Healthcare System and Yale University. It has been granted a waiver of informed consent and is Health Insurance Portability and Accountability Act compliant. This study is reported as per the Strengthening the Reporting of Observational Studies in Epidemiology (STROBE) and reporting of studies conducted using observational routinely collected health data (RECORD) guidelines (see **Supplementary Appendix**).

### Study design and population

We conducted an observational cohort study including all patients aged 18 years or older in active care in the VA between 1 March 2018 and 28 February 2022. We allowed for two years of pre-pandemic follow-up (i.e., 1 March 2018 to 29 February 2020) and two years of pandemic follow-up (i.e., 1 March 2020 to 28 February 2022), covering the same periods of the years to mitigate seasonal variation in mortality trends. Active VA care was defined as the presence of an outpatient or inpatient diagnostic code in the two years prior to each time period, in line with our previous work.^13^ Baseline date was defined as the latest of 1 March 2018 or one year after their first diagnosis code in the two-year period before 1 March 2018, to allow for the recording of baseline covariates. Deaths were ascertained using inpatient records and VA death registry data to capture deaths outside of hospitalisation. Patients were followed until the earliest of date of death, dropped out of care (i.e., 18 months after their last visit), or end of study (i.e., 28 February 2022).

### Covariates

We selected demographic and clinical characteristics that have been evaluated in prior reports as contributors to COVID-19 excess mortality in addition to validated measures of physiologic frailty and comorbidity burden. Demographics included age, sex, race/ethnicity, US census region (i.e., West, South, Midwest, and Northeast), and residence type (i.e., urban, rural). Race and ethnicity were self-reported and categorised as White, Black, Hispanic or Latino (Hispanic), Asian, American Indian/Alaska Native, Pacific Islander/Native Hawaiian, and people of mixed race. In line with previous work,^14^ patients who reported Hispanic ethnicity were included in the Hispanic group regardless of any other self-reported race. Residence type was defined using geographic information system coding based upon established criteria.^15^

The Veterans Aging Cohort Study Index (VACS Index) assesses physiologic frailty by calculating a summary score using a validated algorithm incorporating haemoglobin, alanine transaminase, aspartate transaminase, platelets, creatinine, hepatitis C status, albumin, white blood cell count, body mass index, and age.^16^ The VACS Index is a validated and generalisable risk index that has been shown to predict and discriminate risk of morbidity and mortality in multiple settings.^17,18^ The Charlson Comorbidity Index (CCI) has been a mainstay measure of overall comorbidity burden for decades and is based on diagnostic codes across 17 clinical domains.^19,20^ We examined CCI as a summary score based on established methods as well as individual components in subgroup analyses. Both VACS Index and CCI were ascertained using the most recent laboratory measures and all diagnostic codes that were recorded in the two years prior to baseline, and the status was time-updated on 1 March 2020 using data from the two years prior to 1 March 2020. Patients could therefore be categorised in more than one CCI domain, and these classifications may differ between the pre-pandemic and pandemic periods.

### Statistical analysis

First, we estimated the hazard of mortality during the pandemic period relative to the pre-pandemic period, adjusting for individual-level characteristics. We fit a Cox proportional hazards model with age as the underlying timescale, and the main pandemic exposure variable defined as a time-updated covariate taking the value ‘0’ before 1 March 2020 and ‘1’ from 1 March 2020.

Cox models estimated excess mortality adjusting the baseline hazard for (i) age only and (ii) additionally adjusting for demographic characteristics, VACS Index, and CCI. Only race/ethnicity (5%) and VACS Index (26%) suffered from missing data. We included a missing category for these covariates under the assumption that associations between fully observed covariates and calendar time did not differ across missingness patterns, which would result in unbiased estimates.^21,22^ More details, including the specification of the fully-adjusted Cox model, can be found in eMethods in the **Supplementary Appendix**.

In subgroup analyses, we estimated excess mortality rates, number of excess deaths, and pandemic hazard ratios measuring relative increases in mortality comparing pandemic to pre-pandemic follow-up within each demographic and clinical characteristic. Cox models were specified as the model described above but with the addition of an interaction term between the pandemic time binary indicator and the given characteristic. A separate model was fit for each characteristic.

In secondary analyses, we repeated the analysis above for each of the 17 clinical domains of the CCI by fitting a separate Cox model with an interaction between the pandemic time variable and a binary indicator denoting presence or absence of a diagnostic code within the relevant clinical domain. These models were not adjusted for CCI summary score to mitigate potential issues with collinearity between the CCI summary score and its individual components. Finally, we reran all models, censoring patients at the date of first recorded SARS-CoV-2 infection or COVID-19 diagnosis to understand the extent to which excess mortality could be attributed to COVID-19 versus all other causes. We used the national VA COVID-19 Shared Data Resource, encompassing verified data on all VA patients who had received a laboratory-confirmed diagnosis of SARS-CoV-2 infection as well as cases who were tested external to the VA with a VA clinical note substantiating the diagnosis. We used Microsoft SQL Server Management Studio v18.11 for data management and SAS Enterprise Guide version 8.3 (SAS Institute, Cary, NC) for statistical analyses.

## RESULTS

### Population characteristics

A total of 5,905,747 patients were eligible for follow-up during the pre-pandemic period. Most patients (n=5,488,957; 92.9%) continued follow-up during the pandemic period, while 416,790 (7.1%) dropped out of care or died during the pre-pandemic period. Of the 5,905,747 patients followed in the pre-pandemic period, median age was 65.8 years (interquartile range 51.0-72.9), 91.4% were men, 68.1% were White, 17.1% were Black, and 6.4% were Hispanic; Table 1).

**Table 1.** Population characteristics

|  | In care during pre-pandemic period |  | In care during pandemic period |  |
| --- | --- | --- | --- | --- |
|  | N | (col %) | N | (col %) |
| Number in care, n (%) | 5,905,747 | (100.0) | 5,488,957 | (100.0) |
| Age, years |  |  |  |  |
| 18-44 | 1,068,852 | (18.1) | 948,482 | (17.3) |
| 45-64 | 1,779,684 | (30.1) | 1,598,729 | (29.1) |
| 65-74 | 1,856,101 | (31.4) | 1,711,804 | (31.2) |
| 75-84 | 775,289 | (13.1) | 810,035 | (14.8) |
| ≥85 | 425,821 | (7.2) | 419,907 | (7.7) |
| Sex |  |  |  |  |
| Women | 505,725 | (8.6) | 492,389 | (9.0) |
| Men | 5,400,022 | (91.4) | 4,996,568 | (91.0) |
| Race/ethnicity |  |  |  |  |
| White | 4,019,281 | (68.1) | 3,708,945 | (67.6) |
| Black | 1,010,252 | (17.1) | 960,865 | (17.5) |
| Hispanic | 380,526 | (6.4) | 362,963 | (6.6) |
| Asian | 62,005 | (1.0) | 59,505 | (1.1) |
| AI/AN | 40,068 | (0.7) | 37,613 | (0.7) |
| PI/NH | 43,242 | (0.7) | 40,617 | (0.7) |
| Mixed race | 45,947 | (0.8) | 43,364 | (0.8) |
| Missing | 304,426 | (5.2) | 275,085 | (5.0) |
| Region |  |  |  |  |
| West | 1,250,453 | (21.2) | 1,164,092 | (21.2) |
| South | 2,614,775 | (44.3) | 2,449,788 | (44.6) |
| Midwest | 1,276,192 | (21.6) | 1,176,417 | (21.4) |
| Northeast | 764,327 | (12.9) | 698,660 | (12.7) |
| Residence type |  |  |  |  |
| Rural | 2,065,668 | (35.0) | 1,917,742 | (34.9) |
| Urban | 3,840,079 | (65.0) | 3,571,215 | (65.1) |
| VACS Index |  |  |  |  |
| 1st quartile | 3,185,621 | (53.9) | 2,886,589 | (52.6) |
| 2nd quartile | 670,522 | (11.4) | 676,597 | (12.3) |
| 3rd quartile | 331,541 | (5.6) | 329,386 | (6.0) |
| 4th quartile | 178,877 | (3.0) | 174,874 | (3.2) |
| Missing | 1,539,186 | (26.1) | 1,421,511 | (25.9) |
| Charlson Comorbidity Index |  |  |  |  |
| 0 | 2,969,223 | (50.3) | 2,654,910 | (48.4) |
| 1 | 1,202,465 | (20.4) | 1,087,001 | (19.8) |
| 2 | 785,898 | (13.3) | 750,777 | (13.7) |
| 3 | 395,732 | (6.7) | 394,276 | (7.2) |
| 4 | 237,171 | (4.0) | 250,466 | (4.6) |
| ≥5 | 315,258 | (5.3) | 351,527 | (6.4) |
| Myocardial infarction | 114,967 | (1.9) | 120,485 | (2.2) |
| Congestive heart failure | 314,548 | (5.3) | 328,581 | (6.0) |
| Peripheral vascular disease | 397,563 | (6.7) | 413,310 | (7.5) |
| Cerebrovascular accident | 328,897 | (5.6) | 337,496 | (6.1) |
| Hemiplegia | 34,837 | (0.6) | 34,246 | (0.6) |
| Dementia | 150,975 | (2.6) | 145,257 | (2.6) |
| COPD | 890,890 | (15.1) | 874,682 | (15.9) |
| Connective tissue disease | 79,307 | (1.3) | 80,422 | (1.5) |
| Peptic ulcer disease | 38,404 | (0.7) | 37,194 | (0.7) |
| Diabetes, uncomplicated | 1,452,090 | (24.6) | 1,402,971 | (25.6) |
| Diabetes, end-organ damage | 615,266 | (10.4) | 634,723 | (11.6) |
| Moderate to severe CKD | 450,381 | (7.6) | 494,276 | (9.0) |
| Liver disease, mild | 301,840 | (5.1) | 296,213 | (5.4) |
| Liver disease, moderate to severe | 21,906 | (0.4) | 21,905 | (0.4) |
| HIV/AIDS | 26,684 | (0.5) | 25,833 | (0.5) |
| Cancer, localised | 421,098 | (7.1) | 435,252 | (7.9) |
| Cancer, metastatic | 33,826 | (0.6) | 36,594 | (0.7) |

Abbreviations: AI/AN, American Indian/Alaska Native; PI/NH, Pacific Islander/Native Hawaiian; VACS, Veterans Aging Cohort Study; COPD, chronic obstructive pulmonary disorder; CKD, chronic kidney disease; HIV, human immunodeficiency virus; AIDS, acquired immune deficiency syndrome

Patients were predominately (44.3%) located in the South, and 65.0% resided in urban settings. Over half (53.9%) of all patients were categorised in the lowest (i.e., least frail) quartile of VACS Index, while 3.0% were in the highest quartile (i.e., most frail). Similarly, half (50.3%) of all patients had a CCI score of 0 indicating no recorded diagnoses across the 17 clinical domains, while 5.3% had a CCI score ≥5. In order of decreasing prevalence, 24.6% were diagnosed with diabetes (uncomplicated), 15.1% were diagnosed with chronic obstructive pulmonary disease, and 10.4% were diagnosed with diabetes (end-organ damage). Similar distributions of patient characteristics were observed among those followed during the pandemic period (**Table 1**).

### Excess mortality

There were 358,664 recorded deaths among patients followed for 11,337,771 person-years (PY) during the pre-pandemic period and 429,289 recorded deaths among those followed for 10,309,181 PYs during the pandemic period, resulting in mortality rates of 31.6 deaths/1000 PY and 41.6 deaths/1000 PY, respectively. Overall, the excess mortality rate was 10.0 deaths/1000 PY, resulting in a total of 103,164 excess deaths. Adjusting for age only, the rate of death during the pandemic period was 27% higher (95% confidence interval [CI] 26%-27%; **Table 2**) than in the pre-pandemic period (‘excess mortality’). Excess mortality was slightly lower at 25% (95% CI 25%-26%) after additionally adjusting for demographic characteristics, physiologic frailty, and comorbidity burden.

**Table 2.** Excess mortality estimates adjusted for age, demographic, and clinical characteristics, with and without censoring of COVID-19 follow-up

|  | **Age-adjusted** | **Fully-adjusted** | **Censoring COVID-19** |
| --- | --- | --- | --- |
|  | **HR (95% CI)** | **HR (95% CI)** | **follow-up** |
|  |  |  | **HR (95% CI)** |
| Period |  |  |  |
| Pre-pandemic | 1.00 (ref) | 1.00 (ref) | 1.00 (ref) |
| Pandemic | 1.27 (1.26-1.27) | 1.25 (1.25-1.26) | 1.19 (1.19-1.20) |
| Sex |  |  |  |
| Men |  | 1.00 (ref) | 1.00 (ref) |
| Women |  | 0.65 (0.64-0.66) | 0.65 (0.65-0.67) |
| Race/ethnicity |  |  |  |
| White |  | 1.00 (ref) | 1.00 (ref) |
| Black |  | 0.86 (0.85-0.87) | 0.85 (0.84-0.86) |
| Hispanic |  | 0.75 (0.75-0.76) | 0.74 (0.73-0.75) |
| Asian |  | 0.62 (0.61-0.65) | 0.62 (0.60-0.64) |
| AI/AN |  | 1.11 (1.08-1.14) | 1.09 (1.06-1.13) |
| PI/NH |  | 0.91 (0.89-0.94) | 0.90 (0.87-0.93) |
| Mixed race |  | 0.96 (0.94-0.99) | 0.96 (0.93-0.99) |
| Missing |  | 1.07 (1.06-1.08) | 1.08 (1.07-1.09) |
| Region |  |  |  |
| South |  | 1.00 (ref) | 1.00 (ref) |
| Midwest |  | 0.96 (0.96-0.97) | 0.96 (0.96-0.97) |
| Northeast |  | 0.93 (0.93-0.94) | 0.93 (0.92-0.94) |
| West |  | 1.01 (1.00-1.01) | 1.00 (1.00-1.01) |
| Residence type |  |  |  |
| Rural |  | 1.00 (ref) | 1.00 (ref) |
| Urban |  | 0.98 (0.98-0.99) | 0.98 (0.98-0.99) |
| VACS Index |  |  |  |
| 1st quartile |  | 1.00 (ref) | 1.00 (ref) |
| 2nd quartile |  | 1.93 (1.91-1.95) | 1.95 (1.93-1.97) |
| 3rd quartile |  | 2.96 (2.93-2.98) | 3.01 (2.98-3.03) |
| 4th quartile |  | 4.95 (4.91-5.00) | 5.06 (5.01-5.10) |
| Missing |  | 2.23 (2.21-2.25) | 2.28 (2.26-2.30) |
| Charlson Comorbidity Index |  |  |  |
| 0 |  | 1.00 (ref) | 1.00 (ref) |
| 1 |  | 1.63 (1.62-1.65) | 1.63 (1.62-1.64) |
| 2 |  | 1.84 (1.83-1.85) | 1.83 (1.81-1.84) |
| 3 |  | 2.36 (2.34-2.38) | 2.34 (2.32-2.36) |
| 4 |  | 2.57 (2.55-2.59) | 2.54 (2.52-2.57) |
| ≥5 |  | 3.87 (3.84-3.90) | 3.82 (3.79-3.85) |

*Abbreviations:* COVID-19, coronavirus disease 19; AI/AN, American Indian/Alaska Native; PI/NH, Pacific Islander/Native Hawaiian; HR, hazard ratio; CI, confidence interval; VACS, Veterans Aging Cohort Study
*Notes:* The VACS Index assesses physiologic frailty using a previously validated algorithm. The Charlson Comorbidity Index is a measure of overall comorbidity burden based on diagnostic codes across 17 clinical domains.

### Subgroup analyses

By age group, the rate of excess deaths was highest among patients ≥85 years (44.6 deaths/1000 PY), and the absolute number of excess deaths was highest among patients aged 65 to 74 years (32,909 excess deaths). The relative increase in the hazard of death was highest among patients aged 18 to 44 years (HR 1.33, 95% CI 1.28-1.39), though this group experienced the lowest absolute number of excess deaths (**Figure 1** and **eTable 1**). By race and ethnicity, though the absolute number of excess deaths was greatest among White patients (77,777 excess deaths), the excess mortality rate (14.2 deaths/1000 PY) and pandemic hazard ratio (HR 1.43, 95% CI 1.35-1.52) were highest among American Indian/Alaska Native patients.

**Figure 1.**
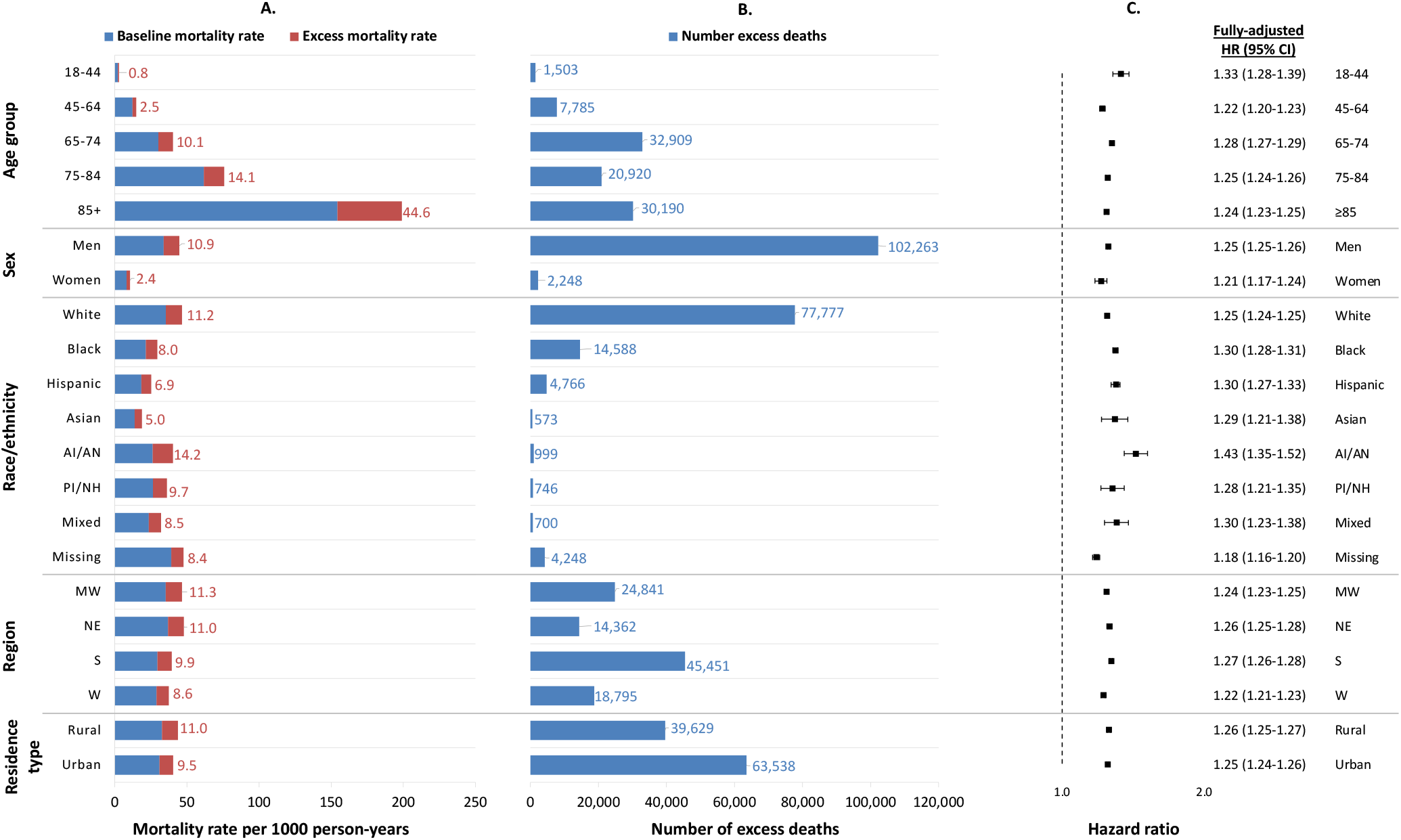
Mortality rates, number of excess deaths, and hazard ratios comparing pre-pandemic and pandemic mortality, by demographic subgroup *Note:* Number of excess deaths are adjusted for the characteristic of interest only. Fully-adjusted hazard ratios were derived from a separate Cox model for each characteristic with an interaction between the pandemic time variable and each given characteristic, and adjusted for all demographics, physiologic frailty, and comorbidity burden. In Panel A, numbers listed refer to excess mortality rate. *Abbreviations:* HR, hazard ratio; CI, confidence interval; AI/AN, American Indian/Alaska Native; PI/NH, Pacific Islander/Native Hawaiian; MW, Midwest; NE, Northeast; S, South; W, West

Excess mortality rate (52.0 deaths/1000 PY) was highest among the most frail patients (fourth quartile of physiologic frailty as measured by the VACS Index), while the highest absolute number of excess deaths (22,253 excess deaths) and largest relative increase in mortality (HR 1.31, 95% CI 1.30-1.32) was observed among the least frail patients (**Figure 2**). Similarly, while the excess mortality rate (16.3 deaths/1000 PY) was highest among patients with the highest comorbidity burden (CCI score ≥5), the highest absolute number of excess deaths (28,931 excess deaths) and relative increase in mortality (HR 1.44, 95% CI 1.43-1.46) were observed among patients with the lowest comorbidity burden (CCI score of 0).

**Figure 2.**
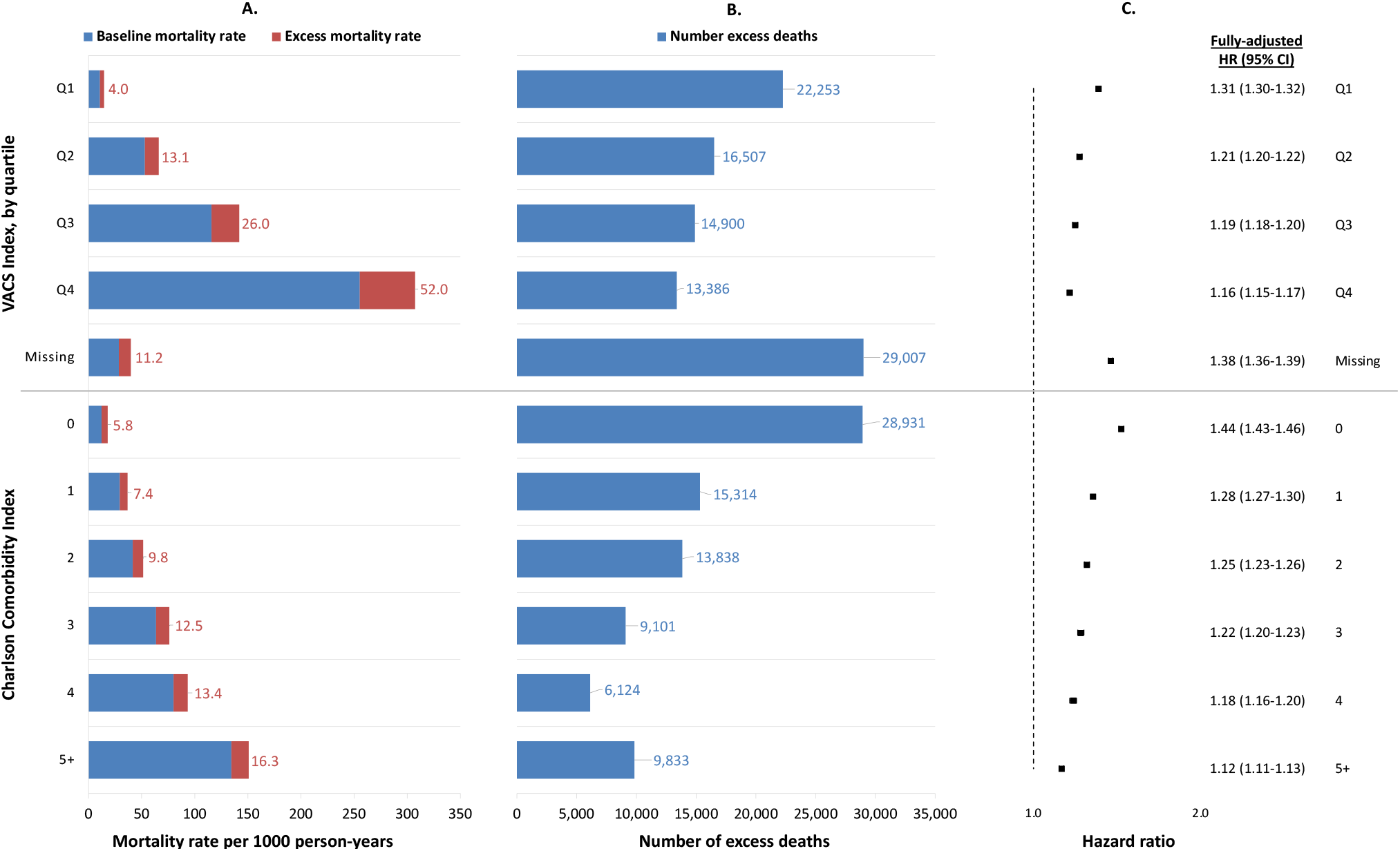
Mortality rates, number of excess deaths, and hazard ratios comparing pre-pandemic and pandemic mortality, by physiologic frailty and comorbidity burden *Note:* Number of excess deaths are adjusted for the characteristic of interest only. Fully-adjusted hazard ratios were derived from a separate Cox model for each characteristic with an interaction between the pandemic time variable and each given characteristic, and adjusted for all demographics, physiologic frailty, and comorbidity burden. In Panel A, numbers listed refer to excess mortality rate. *Abbreviations:* HR, hazard ratio; CI, confidence interval; VACS, Veterans Aging Cohort Study; Q1, first quartile; Q2, second quartile; Q3, third quartile; Q4 fourth quartile; CCI, Charlson Comorbidity Index

### Secondary analyses

Patients with dementia had the highest excess mortality rate (52.2 deaths/1000 PY) and highest relative increase in mortality (HR 1.32, 95% CI 1.30-1.33; **Figure 3** and **eTable 2**). However, patients with diabetes had the highest number of excess deaths, including 36,278 excess deaths among those with uncomplicated diabetes and 21,365 excess deaths among those with diabetes-associated end-organ damage. There were 626,973 (11.4%) patients who had evidence of SARS-CoV-2/COVID-19 during the first two years of the pandemic. After censoring COVID-19 follow-up, the pandemic hazard ratio attenuated from 1.25 (95% CI 1.25-1.26) to 1.19 (95% CI 1.19-1.20) (**Table 1**). Changes in the pandemic hazard ratio after censoring COVID-19 follow-up followed a similar pattern for all demographic and clinical subgroups, with the largest absolute differences observed among Hispanic patients (HR 1.30, 95% CI 1.27-1.33 before censoring and HR 1.19, 95% CI 1.17-1.22 after censoring) and those with the lowest VACS Index (HR 1.31, 95% CI 1.30-1.32 before censoring and HR 1.21, 95% CI 1.20-1.22 after censoring; **eTable 1**).

**Figure 3.**
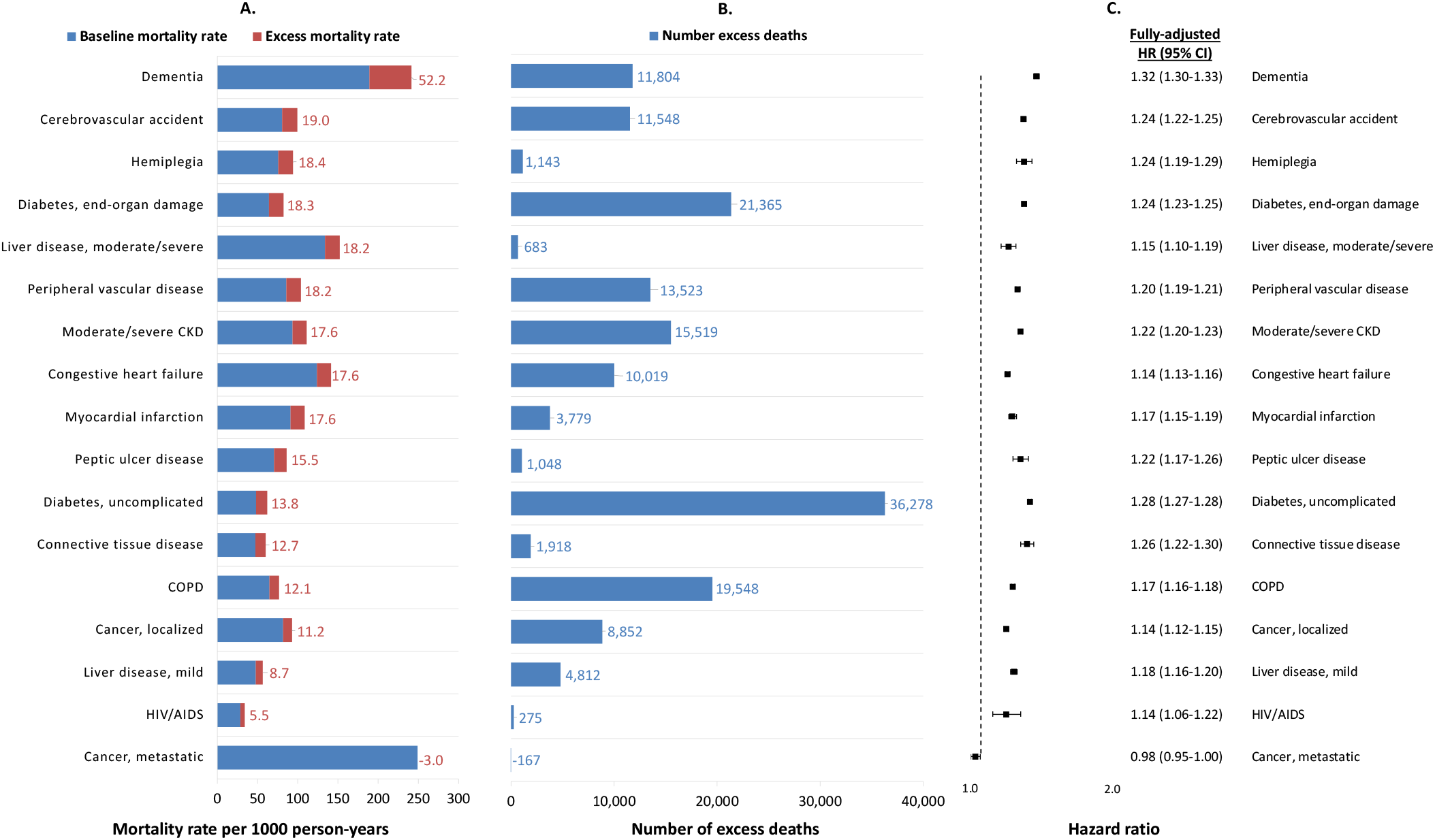
Mortality rates, number of excess deaths, and hazard ratios comparing pre-pandemic and pandemic mortality, by clinical domain (ordered by excess mortality rate) *Note:* Number of excess deaths are adjusted for the characteristic of interest only. Patients can contribute to more than one clinical domain. Fully-adjusted hazard ratios were derived from a separate Cox model for each clinical domain with an interaction between the pandemic time variable and a binary indicator denoting presence or absence of a diagnostic code within the relevant clinical domain, and adjusted for all demographic characteristics and physiologic frailty. In Panel A, numbers listed refer to excess mortality rate. *Abbreviations:* HR, hazard ratio; CI, confidence interval; CKD, chronic kidney disease; COPD, chronic obstructive pulmonary disease; HIV, human immunodeficiency virus; AIDS, acquired immune deficiency syndrome

## DISCUSSION

The impact of the COVID-19 pandemic on overall rates of mortality has been well documented; however, previous work has largely relied on aggregate population-level data. Using individual-level electronic health record data from the largest integrated healthcare system in the US, we demonstrated that the absolute impact as measured by excess mortality rates was typically greatest in groups where the baseline rate of mortality was higher; namely in older age groups and among those with more comorbidities and higher levels of physiologic frailty. However, relative increases in the hazard of mortality during the pandemic were typically greatest among younger age groups and among those with lower physiologic frailty and fewer comorbidities.

Patients with dementia had both the highest absolute excess mortality rate and highest relative increase in mortality. Estimates of excess mortality attenuated but remained elevated after censoring follow-up at first documented SARS-CoV-2 infection or COVID-19, suggesting that factors beyond SARS-CoV-2 infection contributed to the observed excess mortality during the pandemic.

The present analysis adds a unique contribution in the use of individual-level data to estimate and interpret rates of excess mortality associated with the COVID-19 pandemic. Most prior analyses of patterns of excess mortality in the United States have used aggregate data.^1,4,5,23–25^ In some analyses, deaths have been disaggregated by demographic characteristics, including age, sex, race/ethnicity, and region; however, this work has been limited in its ability to adjust for underlying health status. As demonstrated in our previous publication,^26^ estimates of excess mortality from ecological models with aggregate data and survival models with individual-level data yielded nearly identical estimates when adjusting for the same demographic factors. In the present study, we extended our previous report and found that estimates of excess mortality only modestly attenuated from 27% to 25% after additionally accounting for validated, time-updated measures of physiologic frailty and comorbidity burden. Another distinguishing feature of the present study was the availability of verified documentation of laboratory-confirmed SARS-CoV-2 infections and COVID-19 diagnoses at the patient level. We leveraged the available information to censor patients at first evidence of infection thereby appropriately allowing observed follow-up time without infection to contribute to the analysis. However, this approach is susceptible to the challenges of complete recording of SARS-CoV-2 infections and COVID-19 diagnoses, particularly early in the pandemic before case definitions were standardised and testing was available on a widespread basis. Estimates of excess mortality attenuated from 25% to 19% after implementing this additional censoring, suggesting that factors beyond SARS-CoV-2 infection contributed to the observed excess mortality during the pandemic.

The present study provides a systems-level summary of the overall burden of excess mortality during the pandemic in a national integrated healthcare system. Our findings are consistent with previous reports that demonstrated that the highest excess mortality on an absolute scale was among older patients and those who were more frail or had higher comorbidity burden.^27^ However, these groups were observed to have the lowest excess mortality on a relative scale, likely because the baseline rate of death was already high in these groups and there are many competing causes of death. In addition, our calculations of the number of excess deaths, which incorporates the size of each subgroup, suggested that the largest aggregate burden was among patients aged 65 to 74 years and those who were least frail or had no recorded comorbidity. Our findings strongly suggest each of these metrics are important and offer a different story in terms of the impact of COVID-19 on excess mortality in the VA. Studies estimating excess mortality should present findings on both the absolute and relative scales to enable policymakers and operations managers to determine where to allocate resources as we emerge from the pandemic and in future similar outbreaks. The lower relative increases in excess mortality among more frail groups and those with more comorbidities has important implications, and suggests that forward mortality displacement might be less of a phenomenon during the post-pandemic period than had been proposed.^28^

While the analysis of the CCI summary score indicated that those with more comorbidities had higher excess mortality on the absolute scale and lower excess mortality on the relative scale, there were some important clinical subgroups that did not follow this pattern. Notably, patients with dementia had the highest excess mortality on both absolute and relative scales, which was previously identified as an important risk group in other healthcare systems, such as that in the United Kingdom.^29^ Interestingly, patients with metastatic cancer, who are likely to be at greater risk of SARS-CoV-2 infection or severe COVID-19,^30^ appeared to have no excess mortality during the first two years of the pandemic. These findings likely underscore the importance of distinguishing infection risk from mortality risk once infected. Patients with dementia are more likely to reside in nursing homes, making them more likely to acquire SARS-CoV-2 infection in addition to their higher mortality risk once infected.^31^ Patients with metastatic cancer were identified as an at-risk group and instructed to shelter at home or take great precaution in public spaces, which may have reduced their probability of infection.

Although there are many published reports highlighting racial and ethnic disparities in testing positive for SARS-CoV-2,^14,32^ we have previously shown that there are no racial or ethnic disparities in the probability of 30-day mortality among those who tested positive in the VA.^13^ While comprising only 1% of the present study, American Indian/Alaska Native patients experienced the largest absolute and relative increases in mortality during the pandemic, highlighting the need for more focused assessment and evidence-based interventions in partnership with affected racial and ethnic minority communities.

This study elucidated patterns of excess mortality associated with the COVID-19 pandemic leveraging individual-level data on demographics and clinical characteristics from a national healthcare system. Study strengths included the ability to adjust estimates of excess mortality for underlying health status and compare magnitude of excess mortality within levels of physiologic frailty and comorbidity burden. This study also has some limitations. First, this study included Veterans currently receiving care in the VA, who are older and have a higher prevalence of chronic health conditions than the general US population.^33,34^ Prior research has established that after adjusting for age, sex, race/ethnicity, region, and rural/urban residence, all of which were included in this study, there is no difference in total disease burden between Veterans and non-Veterans.^35^ While we expect absolute measures to differ across the population under study, relative measures in the present study are more likely to generalise to the general US population, which we have demonstrated in previous work.^26^ Second, while most variables used in the present study were complete, nearly one in four patients had missing labs to calculate the VACS Index and were categorised separately. Third, although secondary analyses utilised records of laboratory-confirmed SARS-CoV-2 infections and COVID-19 diagnoses from both VA and external sources, some patients may be misclassified if they tested positive elsewhere and did not self-report the diagnoses at a subsequent VA visit. The US Centers for Disease Control and Prevention (CDC) nationwide seroprevalence study estimated that cumulative reported COVID-19 cases was 14.9% and infection-induced seroprevalence was 28.8% in December 2021.^36,37^ Among patients in the present study, 11.4% had a record of a SARS-CoV-2 infection or COVID-19 diagnosis by February 2022. Patients with less severe infections identified through home testing as well as those who died from severe infections in non-VA hospitals are both likely to be misclassified, which would influence our results of the secondary analyses in both directions.

In conclusion, we report important differences in patterns of excess mortality between clinically-defined risk groups. The relative increase in mortality was smaller in groups with higher frailty and comorbidity burden. However, because baseline death rates were higher in those groups, the absolute increase in mortality rates was greatest among these higher risk groups. The use of individual-level data provided important clinical context for patterns of excess mortality in the United States during the COVID-19 pandemic.

## Data Availability

Due to US Department of Veterans Affairs (VA) regulations and our ethics agreements, the analytic data sets used for this study are not permitted to leave the VA firewall without a data use agreement. This limitation is consistent with other studies based on VA data. However, VA data are made freely available to researchers with an approved VA study protocol. For more information, please visit https://www.virec.research.va.gov or contact the VA Information Resource Center at.

https://github.com/VA-CareDisruptions/VA-Individual-Excess-Mortality

## Ethics approval

This study was approved by the institutional review boards of Yale University (ref #1506016006) and VA Connecticut Healthcare System (ref #AJ0013). It has been granted a waiver of informed consent and is compliant with the Health Insurance Portability and Accountability Act.

## Author contributions

ACJ and CTR conceived the study. CTR curated the data. CTR performed the formal analysis. AV, LD, and ACJ acquired funding. DMW, KB, ACJ, and CTR designed the methodology. ACJ and CTR managed and coordinated the project. ACJ procured resources to carry out the study.

CR developed programming. ACJ and CTR provided oversight and leadership of the project. DMW and CTR prepared data visualisations. DMW and CTR wrote the first draft of the manuscript. All the authors wrote (reviewed and edited) the manuscript. The senior author attests that all listed authors meet authorship criteria and that no others meeting the criteria have been omitted.

## Acknowledgements

This work uses data provided by patients and collected by the VA as part of their care and support. The views and opinions expressed in this manuscript are those of the authors and do not necessarily represent those of the Department of Veterans Affairs or the United States government.

