## Supplementary Appendix for "Absolute and relative excess mortality across demographic and clinical subgroups during the COVID-19 pandemic: an individual-level cohort study from a nationwide healthcare system of US Veterans"

### eMethods

To create the time-updated pandemic exposure variable, we performed Lexis expansion based on calendar time at 1 March 2020, thereby creating up to two rows of data per patient each capturing follow-up observed before and after 1 March 2020.^1^ After Lexis expansion, we created a binary indicator set to 0 for records capturing follow-up in the pre-pandemic period and 1 for records capturing follow-up in the pandemic period, which was used as the primary exposure variable in Cox proportional hazards models. We used age as the underlying timescale in the Cox models (i.e., patients entered the analysis at their baseline age and exited at their event/censoring age) to allow for a completely non-parametric accounting of age, which is preferred when age is associated with the outcome in a non-linear fashion as it is with mortality.^2^

The fully-adjusted Cox model was specified as follows:

$$h\left( a | X \right)= h_{0}\left( a \right)*exp(\beta_{1}*Pandemic + \gamma_{1}*Sex + \Sigma_{i}^{7}\delta_{i}*{Race/ethnicity}_{i} + \Sigma_{j}^{3}\epsilon_{j}*{Region}_{j}+ \zeta_{1}*Residence\_type + \Sigma_{k}^{4}\eta_{k}*{Frailty}_{k}\left( a \right)+ \Sigma_{l}^{5}\theta_{l}*{Comorbidity}_{l}\left( a \right))$$

Where:

- $h\left( a | X \right)$ is the hazard function at age $a$, given the covariates $X$
- $h_{0}\left( a \right)$ is the baseline hazard function at a specific age $a$
- Fixed effects included pandemic period, sex, race/ethnicity, region, and residence type
- Physiologic frailty as measured by the Veterans Aging Cohort Study Index and comorbidity burden as measured by the Charlson Comorbidity Index were time-updated at a specific age $a$

In the subgroup analyses, excess mortality rates were estimated by subtracting the rate of mortality in the pandemic period from the rate of mortality in the pre-pandemic period. Number of excess deaths were estimated by multiplying the rate of mortality in the pre-pandemic period by the total number of person-years observed in the pandemic period to obtain expected numbers of deaths, and took the difference between expected and observed deaths in the pandemic period. We repeated this exercise within each category of the included demographic and clinical characteristics.

| eTable 1. Mortality rates, number of excess deaths, and hazard ratios comparing pre-pandemic and pandemic mortality, by demographic subgroup, physiologic frailty, and comorbidity burden, with and without censoring of COVID-19 follow-up | | | | | | |
| --- | --- | --- | --- | --- | --- | --- |
|  | **Baseline mortality rate** | **Excess mortality rate** | **Number excess deaths** | **Age-adjusted HR (95% CI)** | **Fully-adjusted HR (95% CI)** | **Censoring COVID-19 follow-up HR (95% CI)** |
| Overall | 31.63 | 10.01 | 103,164 | 1.27 (1.26-1.27) | 1.25 (1.25-1.26) | 1.19 (1.19-1.20) |
| Age |  |  |  |  |  |  |
| 18-44 | 2.14 | 0.83 | 1,503 | 1.34 (1.29-1.40) | 1.33 (1.28-1.39) | 1.27 (1.22-1.32) |
| 45-64 | 12.30 | 2.52 | 7,785 | 1.19 (1.18-1.21) | 1.22 (1.20-1.23) | 1.15 (1.13-1.16) |
| 65-74 | 30.19 | 10.13 | 32,909 | 1.29 (1.28-1.30) | 1.28 (1.27-1.29) | 1.19 (1.18-1.20) |
| 75-84 | 61.76 | 14.13 | 20,920 | 1.28 (1.27-1.29) | 1.25 (1.24-1.26) | 1.19 (1.18-1.20) |
| 85+ | 154.35 | 44.59 | 30,190 | 1.26 (1.25-1.27) | 1.24 (1.23-1.25) | 1.20 (1.19-1.21) |
| Sex |  |  |  |  |  |  |
| Men | 33.87 | 10.93 | 102,263 | 1.27 (1.27-1.28) | 1.25 (1.25-1.26) | 1.19 (1.19-1.20) |
| Women | 8.23 | 2.36 | 2,248 | 1.21 (1.17-1.24) | 1.21 (1.17-1.24) | 1.15 (1.12-1.18) |
| Race/ethnicity |  |  |  |  |  |  |
| White | 35.40 | 11.21 | 77,777 | 1.27 (1.26-1.27) | 1.25 (1.24-1.25) | 1.19 (1.18-1.20) |
| Black | 21.53 | 7.96 | 14,588 | 1.28 (1.26-1.30) | 1.30 (1.28-1.31) | 1.21 (1.19-1.22) |
| Hispanic | 18.36 | 6.89 | 4,766 | 1.31 (1.28-1.34) | 1.30 (1.27-1.33) | 1.19 (1.17-1.22) |
| Asian | 13.74 | 5.05 | 573 | 1.31 (1.23-1.40) | 1.29 (1.21-1.38) | 1.22 (1.15-1.31) |
| AI/AN | 26.21 | 14.15 | 999 | 1.45 (1.37-1.54) | 1.43 (1.35-1.52) | 1.34 (1.26-1.42) |
| PI/NH | 26.49 | 9.70 | 746 | 1.31 (1.24-1.38) | 1.28 (1.21-1.35) | 1.19 (1.13-1.26) |
| Mixed race | 23.56 | 8.54 | 700 | 1.32 (1.24-1.40) | 1.30 (1.23-1.38) | 1.23 (1.16-1.31) |
| Missing | 39.20 | 8.44 | 4,248 | 1.19 (1.17-1.22) | 1.18 (1.16-1.20) | 1.15 (1.13-1.17) |
| Region |  |  |  |  |  |  |
| Midwest | 35.28 | 11.27 | 24,841 | 1.27 (1.26-1.28) | 1.24 (1.23-1.25) | 1.18 (1.17-1.19) |
| Northeast | 36.90 | 11.02 | 14,362 | 1.26 (1.25-1.28) | 1.26 (1.25-1.28) | 1.20 (1.19-1.22) |
| South | 29.64 | 9.86 | 45,451 | 1.28 (1.27-1.29) | 1.27 (1.26-1.28) | 1.21 (1.20-1.21) |
| West | 28.91 | 8.59 | 18,795 | 1.24 (1.22-1.25) | 1.22 (1.21-1.23) | 1.16 (1.15-1.17) |
| Residence type |  |  |  |  |  |  |
| Rural | 32.76 | 10.99 | 39,629 | 1.27 (1.26-1.28) | 1.26 (1.25-1.27) | 1.20 (1.19-1.21) |
| Urban | 31.03 | 9.48 | 63,538 | 1.26 (1.26-1.27) | 1.25 (1.24-1.26) | 1.19 (1.18-1.19) |
| VACS Index |  |  |  |  |  |  |
| 1st quartile | 10.72 | 3.95 | 22,253 | 1.34 (1.32-1.35) | 1.31 (1.30-1.32) | 1.21 (1.20-1.22) |
| 2nd quartile | 53.03 | 13.05 | 16,507 | 1.25 (1.23-1.26) | 1.21 (1.20-1.22) | 1.14 (1.12-1.15) |
| 3rd quartile | 115.83 | 26.03 | 14,900 | 1.23 (1.22-1.25) | 1.19 (1.18-1.20) | 1.13 (1.12-1.14) |
| 4th quartile | 255.35 | 51.98 | 13,386 | 1.21 (1.20-1.22) | 1.16 (1.15-1.17) | 1.12 (1.10-1.13) |
| Missing | 28.56 | 11.24 | 29,007 | 1.38 (1.37-1.39) | 1.38 (1.36-1.39) | 1.34 (1.33-1.35) |
| Charlson Comorbidity Index | |  |  |  |  |  |
| 0 | 12.31 | 5.76 | 28,931 | 1.42 (1.40-1.43) | 1.44 (1.43-1.46) | 1.39 (1.38-1.41) |
| 1 | 29.41 | 7.38 | 15,314 | 1.25 (1.23-1.26) | 1.28 (1.27-1.30) | 1.23 (1.21-1.24) |
| 2 | 41.69 | 9.75 | 13,838 | 1.22 (1.20-1.23) | 1.25 (1.23-1.26) | 1.18 (1.17-1.20) |
| 3 | 63.59 | 12.48 | 9,101 | 1.18 (1.17-1.20) | 1.22 (1.20-1.23) | 1.15 (1.14-1.16) |
| 4 | 80.01 | 13.43 | 6,124 | 1.15 (1.13-1.16) | 1.18 (1.16-1.20) | 1.11 (1.09-1.13) |
| ≥5 | 134.50 | 16.28 | 9,833 | 1.09 (1.08-1.10) | 1.12 (1.11-1.13) | 1.05 (1.04-1.06) |
| *Notes:* Number of excess deaths are adjusted for the characteristic of interest only. Fully-adjusted hazard ratios were derived from a separate Cox model for each characteristic with an interaction between the pandemic time variable and each given characteristic, and adjusted for all demographics, physiologic frailty, and comorbidity burden. | | | | | | |
| Abbreviations: HR, hazard ratio; CI, confidence interval; AI/AN, American Indian/Alaska Native; PI/NH, Pacific Islander/Native Hawaiian; MW, Midwest; NE, Northeast; S, South; W, West; VACS, Veterans Aging Cohort Study | | | | | | |

| eTable 2. Mortality rates, number of excess deaths, and hazard ratios comparing pre-pandemic and pandemic mortality, by clinical domain (ordered by excess mortality rate), with and without censoring of COVID-19 follow-up | | | | | | |
| --- | --- | --- | --- | --- | --- | --- |
|  | **Baseline mortality rate** | **Excess mortality rate** | **Number excess deaths** | **Age-adjusted HR (95% CI)** | **Fully-adjusted HR (95% CI)** | **Censoring COVID-19 follow-up HR (95% CI)** |
| Dementia | 189.22 | 52.21 | 11,804 | 1.30 (1.28-1.32) | 1.32 (1.30-1.33) | 1.24 (1.23-1.26) |
| Cerebrovascular accident | 80.60 | 18.98 | 11,548 | 1.22 (1.20-1.23) | 1.24 (1.22-1.25) | 1.16 (1.14-1.17) |
| Hemiplegia | 75.74 | 18.41 | 1,143 | 1.20 (1.15-1.25) | 1.24 (1.19-1.29) | 1.16 (1.12-1.21) |
| Diabetes, end-organ damage | 64.11 | 18.30 | 21,365 | 1.23 (1.22-1.24) | 1.24 (1.23-1.25) | 1.15 (1.14-1.16) |
| Liver disease, moderate/severe | 133.94 | 18.22 | 683 | 1.06 (1.02-1.10) | 1.15 (1.10-1.19) | 1.09 (1.05-1.13) |
| Peripheral vascular disease | 85.82 | 18.18 | 13,523 | 1.18 (1.17-1.20) | 1.20 (1.19-1.21) | 1.12 (1.11-1.14) |
| Moderate/severe CKD | 93.54 | 17.59 | 15,519 | 1.19 (1.18-1.20) | 1.22 (1.20-1.23) | 1.14 (1.13-1.15) |
| Congestive heart failure | 123.81 | 17.58 | 10,019 | 1.12 (1.11-1.14) | 1.14 (1.13-1.16) | 1.08 (1.06-1.09) |
| Myocardial infarction | 91.08 | 17.56 | 3,779 | 1.16 (1.14-1.18) | 1.17 (1.15-1.19) | 1.09 (1.07-1.11) |
| Peptic ulcer disease | 70.65 | 15.46 | 1,048 | 1.19 (1.14-1.23) | 1.22 (1.17-1.26) | 1.14 (1.10-1.19) |
| Diabetes, uncomplicated | 48.19 | 13.82 | 36,278 | 1.26 (1.25-1.27) | 1.28 (1.27-1.28) | 1.19 (1.18-1.20) |
| Connective tissue disease | 47.36 | 12.72 | 1,918 | 1.24 (1.20-1.28) | 1.26 (1.22-1.30) | 1.16 (1.13-1.20) |
| COPD | 64.70 | 12.12 | 19,548 | 1.15 (1.14-1.16) | 1.17 (1.16-1.18) | 1.11 (1.10-1.12) |
| Cancer, localized | 81.78 | 11.22 | 8,852 | 1.12 (1.11-1.13) | 1.14 (1.12-1.15) | 1.08 (1.07-1.09) |
| Liver disease, mild | 47.86 | 8.65 | 4,812 | 1.10 (1.09-1.12) | 1.18 (1.16-1.20) | 1.11 (1.09-1.13) |
| HIV/AIDS | 28.56 | 5.55 | 275 | 1.09 (1.01-1.17) | 1.14 (1.06-1.22) | 1.08 (1.00-1.16) |
| Cancer, metastatic | 249.09 | -3.00 | -167 | 0.95 (0.93-0.98) | 0.98 (0.95-1.00) | 0.95 (0.93-0.98) |
| *Note:* Number of excess deaths are adjusted for the characteristic of interest only. Patients can contribute to more than one clinical domain. Fully-adjusted hazard ratios were derived from a separate Cox model for each clinical domain with an interaction between the pandemic time variable and a binary indicator denoting presence or absence of a diagnostic code within the relevant clinical domain, and adjusted for all demographic characteristics and physiologic frailty. | | | | | | |
| Abbreviations: HR, hazard ratio; CI, confidence interval; CKD, chronic kidney disease; COPD, chronic obstructive pulmonary disease; HIV, human immunodeficiency virus; AIDS, acquired immune deficiency syndrome | | | | | | |

### STROBE/RECORD statement

|  | **Item No.** | **STROBE items** | **Location in manuscript where items are reported** | **RECORD items** | **Location in manuscript where items are reported** |
| --- | --- | --- | --- | --- | --- |
| **Title and abstract** | | | | | |
|  | 1 | (a) Indicate the study’s design with a commonly used term in the title or the abstract (b) Provide in the abstract an informative and balanced summary of what was done and what was found | (a) Title & Abstract  (b) Abstract | RECORD 1.1: The type of data used should be specified in the title or abstract. When possible, the name of the databases used should be included.  RECORD 1.2: If applicable, the geographic region and timeframe within which the study took place should be reported in the title or abstract.  RECORD 1.3: If linkage between databases was conducted for the study, this should be clearly stated in the title or abstract. | 1.1: Title & Abstract  1.2: Title & Abstract  1.3: Abstract |
| **Introduction** | | | | | |
| Background rationale | 2 | Explain the scientific background and rationale for the investigation being reported | Introduction (Para 1 & 2) |  |  |
| Objectives | 3 | State specific objectives, including any prespecified hypotheses | Introduction (End of Para 2) |  |  |
| **Methods** | | | | | |
| Study Design | 4 | Present key elements of study design early in the paper | Methods (Study design and population) |  |  |
| Setting | 5 | Describe the setting, locations, and relevant dates, including periods of recruitment, exposure, follow-up, and data collection | Methods (Data source; Study design and population; Covariates; Statistical analysis) |  |  |
| Participants | 6 | *(a) Cohort study* - Give the eligibility criteria, and the sources and methods of selection of participants. Describe methods of follow-up  *Case-control study* - Give the eligibility criteria, and the sources and methods of case ascertainment and control selection. Give the rationale for the choice of cases and controls  *Cross-sectional study* - Give the eligibility criteria, and the sources and methods of selection of participants  *(b) Cohort study* - For matched studies, give matching criteria and number of exposed and unexposed  *Case-control study* - For matched studies, give matching criteria and the number of controls per case | (a) Methods (Study design and population; Statistical analysis) | RECORD 6.1: The methods of study population selection (such as codes or algorithms used to identify subjects) should be listed in detail. If this is not possible, an explanation should be provided.  RECORD 6.2: Any validation studies of the codes or algorithms used to select the population should be referenced. If validation was conducted for this study and not published elsewhere, detailed methods and results should be provided.  RECORD 6.3: If the study involved linkage of databases, consider use of a flow diagram or other graphical display to demonstrate the data linkage process, including the number of individuals with linked data at each stage. | 6.1: Methods (Study design and population; Statistical analysis)  6.2: Methods (Covariates)  6.3: n/a |
| Variables | 7 | Clearly define all outcomes, exposures, predictors, potential confounders, and effect modifiers. Give diagnostic criteria, if applicable. | Methods (Covariates; Statistical analysis) | RECORD 7.1: A complete list of codes and algorithms used to classify exposures, outcomes, confounders, and effect modifiers should be provided. If these cannot be reported, an explanation should be provided. | 7.1: Methods (Covariates; Statistical analysis) |
| Data sources/ measurement | 8 | For each variable of interest, give sources of data and details of methods of assessment (measurement).  Describe comparability of assessment methods if there is more than one group | Methods (Covariates; Statistical analysis). |  |  |
| Bias | 9 | Describe any efforts to address potential sources of bias | Methods (Covariates; Statistical analysis; Secondary analyses) |  |  |
| Study size | 10 | Explain how the study size was arrived at | Methods (all) |  |  |
| Quantitative variables | 11 | Explain how quantitative variables were handled in the analyses. If applicable, describe which groupings were chosen, and why | Methods (Covariates; Statistical analysis) |  |  |
| Statistical methods | 12 | (a) Describe all statistical methods, including those used to control for confounding  (b) Describe any methods used to examine subgroups and interactions  (c) Explain how missing data were addressed  (d) *Cohort study* - If applicable, explain how loss to follow-up was addressed  *Case-control study* - If applicable, explain how matching of cases and controls was addressed  *Cross-sectional study* - If applicable, describe analytical methods taking account of sampling strategy  (e) Describe any sensitivity analyses | (a-c) Methods (Covariates; Statistical analysis; Secondary analyses)  (d) Methods (Study design and population)  (e) Methods (Secondary analyses) |  |  |
| Data access and cleaning methods |  | .. |  | RECORD 12.1: Authors should describe the extent to which the investigators had access to the database population used to create the study population.  RECORD 12.2: Authors should provide information on the data cleaning methods used in the study. | 12.1: Methods (Role of the funding source)  12.2: Author contributions |
| Linkage |  | .. |  | RECORD 12.3: State whether the study included person-level, institutional-level, or other data linkage across two or more databases. The methods of linkage and methods of linkage quality evaluation should be provided. | Methods (Study design and population; Secondary analyses) |
| **Results** | | | | | |
| Participants | 13 | (a) Report the numbers of individuals at each stage of the study (*e.g.*, numbers potentially eligible, examined for eligibility, confirmed eligible, included in the study, completing follow-up, and analysed)  (b) Give reasons for non-participation at each stage.  (c) Consider use of a flow diagram | (a-b) Results (Population characteristics)  (c) n/a | RECORD 13.1: Describe in detail the selection of the persons included in the study (*i.e.,* study population selection) including filtering based on data quality, data availability and linkage. The selection of included persons can be described in the text and/or by means of the study flow diagram. | 13.1: Results (Population characteristics) |
| Descriptive data | 14 | (a) Give characteristics of study participants (*e.g.*, demographic, clinical, social) and information on exposures and potential confounders  (b) Indicate the number of participants with missing data for each variable of interest  (c) *Cohort study* - summarise follow-up time (*e.g.*, average and total amount) | (a) Results (Population characteristics; Table 1)  (b) Methods (Covariates); Table 1  (c) Results (Age-standardised incidence rates) |  |  |
| Outcome data | 15 | *Cohort study* - Report numbers of outcome events or summary measures over time  *Case-control study* - Report numbers in each exposure category, or summary measures of exposure  *Cross-sectional study* - Report numbers of outcome events or summary measures | Results (Excess mortality) |  |  |
| Main results | 16 | (a) Give unadjusted estimates and, if applicable, confounder-adjusted estimates and their precision (e.g., 95% confidence interval). Make clear which confounders were adjusted for and why they were included  (b) Report category boundaries when continuous variables were categorized  (c) If relevant, consider translating estimates of relative risk into absolute risk for a meaningful time period | (a) Results (Excess mortality; Subgroup analyses; Secondary analyses; Figure 1; Figure 2; Figure 3; eTable 1; eTable 2)  (b) Results (all)  (c) Results (Excess mortality) |  |  |
| Other analyses | 17 | Report other analyses done—e.g., analyses of subgroups and interactions, and sensitivity analyses | Results (Subgroup analyses; Secondary analyses) |  |  |
| **Discussion** | | | | | |
| Key results | 18 | Summarise key results with reference to study objectives | Discussion (para 1) |  |  |
| Limitations | 19 | Discuss limitations of the study, taking into account sources of potential bias or imprecision. Discuss both direction and magnitude of any potential bias | Discussion (para 7) | RECORD 19.1: Discuss the implications of using data that were not created or collected to answer the specific research question(s). Include discussion of misclassification bias, unmeasured confounding, missing data, and changing eligibility over time, as they pertain to the study being reported. | 19.1 Discussion (para 7) |
| Interpretation | 20 | Give a cautious overall interpretation of results considering objectives, limitations, multiplicity of analyses, results from similar studies, and other relevant evidence | Discussion (All) |  |  |
| Generalisability | 21 | Discuss the generalisability (external validity) of the study results | Discussion (para 7) |  |  |
| **Other Information** | | | | | |
| Funding | 22 | Give the source of funding and the role of the funders for the present study and, if applicable, for the original study on which the present article is based | Funding statement |  |  |
| Accessibility of protocol, raw data, and programming code |  | .. |  | RECORD 22.1: Authors should provide information on how to access any supplemental information such as the study protocol, raw data, or programming code. | Data availability statement |
